# Dynamics of anti-Spike IgG antibody titer after the third BNT162b2 COVID-19 vaccination in the Japanese health care workers

**DOI:** 10.1101/2022.04.10.22273678

**Authors:** Hiroaki Ikezaki, Hideyuki Nomura, Nobuyuki Shimono

## Abstract

**Introduction:** Many countries are administering a third dose of some coronavirus disease 2019 (COVID-19) vaccines, but the evaluation of vaccine-induced immunity is insufficient. This study aimed to evaluate anti-spike immunoglobulin G (IgG) titers in the health care workers after the third BNT162b2 vaccination.

**Methods:** Dynamics of anti-spike IgG titers were assessed two months following the third BNT162b2 vaccination in 52 participants. All participants received the primary series of vaccination with BNT162b2 and received the third dose eight months after the second vaccination. Associations between anti-spike IgG titer, baseline characteristics, and adverse reactions were also evaluated.

**Results:** The geometric mean titer of anti-spike IgG one month after the third vaccination was 17400 AU/ml, which increased to approximately 30 times immediately before the third vaccination and approximately twice that one month after the second vaccination. In addition, participants with anti-spike IgG titers less than 10000 AU/ml after the second vaccination tended to have higher increases in ant-spike IgG titers before and after the third vaccination.

The decline rate of anti-spike IgG was significantly slower after the third vaccination as 35.7% than that after the second vaccination as 59.1%. The anti-spike IgG titer was significantly negatively associated with age (*r* = -0.31). Participants who had a headache at the vaccination showed significantly higher anti-spike IgG titer than those without a headache.

**Conclusions:** The anti-spike IgG induced by primary immunization with BNT162b2 waned over time. The third dose of BNT162b2 substantially increased the anti-spike IgG with a slower decline rate.

## Introduction

Although the vaccines against coronavirus disease 2019 (COVID-19), including messenger RNA (mRNA) vaccines, were highly effective in preventing COVID-19 at the beginning of the vaccination initiation [1, 2], it has become increasingly difficult to prevent the spread of COVID-19 with the first series of vaccines as mutant variants have emerged. On November 26, 2021, the World Health Organization named the new severe acute respiratory syndrome coronavirus 2 (SARS-CoV-2) B.1.1.529 variant omicron and classified it as a Variant of Concern based on a rapid increase in confirmed cases of SARS-CoV-2 infection with this variant in South Africa [3, 4]. The omicron variant increases transmissibility and immune evasion even after natural infection and vaccinations because it has a large number of mutations, including multiple mutations in the receptor-binding domain of the spike protein [5]. In addition, early laboratory data indicate that the neutralizing antibody response to the omicron strain is substantially reduced compared to the original strain or the delta variant in vaccinated individuals [6-9]. COVID-19 vaccines are highly effective against both symptomatic and severe diseases caused by the original strain and the alpha variant [1, 2]. Although waning of anti-spike immunoglobulin G (IgG) titer and protection has been observed with time after vaccination, especially with the delta variant, booster (third) doses provide rapid and robust increases in anti-spike IgG titer and protection against both mild and severe diseases [10-12].

Based on these findings, many countries have commenced administering a booster (third) vaccine dose to curb the pandemic [10-13]. In addition, Israel was the first country in the world to administer the fourth dose in December 2021 [14]. The booster doses of both the BNT162b2 and mRNA-1273 have been approved in Japan since December 2021 for those over 12 years old [15]. However, there are a few reports of post-booster anti-spike IgG antibody response and adverse reactions in East Asian populations; therefore, we aimed to evaluate the post-booster anti-spike IgG titer and adverse reactions in the Japanese population.

## Materials and Methods

### Study participants and design

Study participants were recruited from health care workers in Haradoi Hospital, a care-mix hospital in Fukuoka [16, 17]. Of the 485 health care workers in this hospital, 52 (10.7%) participated in this long-term prospective study. Most of the study participants were nurses, and approximately 85% were women. All three vaccines administrated to the participants were BNT162b2 mRNA COVID-19 vaccine (Comirnaty^®^: Pfizer/BioNTech). All participants were offered first, second, and third doses of the vaccine on March, April, and December 2021. All participants provided written informed consent prior to enrollment. This study was carried out in accordance with the principles of the Declaration of Helsinki, as revised in 2008, and approved by the Haradoi hospital institutional ethics review committee prior to data collection (Approval No. 2020-08).

The main objective of this study was to evaluate the dynamics of anti-spike IgG titers and the anti-spike IgG titers were measured nine times; before the first vaccination, three weeks after the first vaccination (just before the second vaccination), one, two, four, and six months after the second vaccination, before the third vaccination (approximately eight months after the second vaccination), and one and two months after the third vaccination. The secondary objective of this study was to assess the safety of BNT162b2 mRNA COVID-19 vaccine by blood tests and interviews of adverse reactions at the vaccinations. Participants provided information on their height, weight, smoking habits (current, past, or never), drinking habits (daily, often, or never), allergies, medical history, medications, whether they had had adverse reactions to the second and third vaccinations (fever, fatigue, headache, and swelling of axillary lymph nodes), and whether they had needed antipyretics.

### Laboratory measurements

Levels of anti-spike IgG were quantified using the SARS-CoV-2 IgG II Quant assay (Abbott Diagnostics, Chicago, IL, USA) [18]. Participants underwent blood testing to quantitatively assess anti-spike IgG nine times (before the first vaccination, just before the second vaccination, one, two, four, and six months after the second vaccination, just before the third vaccination, and one and two months after the third vaccination). The results of anti-spike IgG are expressed as arbitrary units per milliliter (AU/ml) (positive threshold: 50 AU/ml). We also performed IgG / immunoglobulin M (IgM) antibody qualitative tests against the SARS-CoV-2 nucleocapsid protein (positive thresholds: 1.40 index [S/C] for anti-nucleocapsid IgG and 1.00 index [S/C] for anti-nucleocapsid IgM) for all participants to exclude the effects of SARS-CoV-2 infection. Participants also had blood tests for total bilirubin, aspartate aminotransferase (AST), alanine aminotransferase (ALT), γ-glutamyl transpeptidase (γ-GTP), and serum creatinine, using standard enzymatic methods. Estimated glomerular filtration rate was calculated using the following equation: 194 × serum creatinine^-1.094^ × age^-0.287^ (× 0.739 [if women]).

### Statistical analysis

Data are expressed as median values with 25^th^ and 75^th^ percentile values for continuous variables. The geometric mean titers (GMT) of anti-spike IgG were calculated. Categorical variables are reported as frequencies and percentages. The Mann–Whitney U test was used to compare two groups, and the Kruskal–Wallis test was used to compare three groups. The Tukey–Kramer method was used for each two-group comparison among three groups. Anti-spike IgG levels, with age and sex adjustment, were determined by the least means square method. McNemar’s test was used to evaluate for differences between adverse reactions at the second and third vaccinations. The Wilcoxon signed-rank test was used to evaluate for differences in laboratory data before and after the vaccinations. All analyses were performed using SAS version 9.4 (SAS Institute Inc., Cary, NC). A *P* value less than 0.05 was considered statistically significant.

## Results

### Baseline characteristics of the participants

Table 1 shows the baseline characteristics of the 52 participants of this study. The median age was 40.5 years, 84.6% were women, and the median body mass index (BMI) was 20.9 kg/m^2^. There were no current smokers, 13.5% were daily alcohol drinkers, and 17.3% had an allergy. Among the study participants, 25 (48.1%) had comorbidities. One had rheumatoid arthritis but did not take corticosteroid or immunosuppressants. Another had a history of colorectal cancer without any evidence of recurrence at the enrollment of this study. Before the first vaccination, the titers of anti-spike IgG and the anti-nucleocapsid IgG and IgM of all 49 participants were below the positive threshold, indicating no one had had COVID-19 before participating in this study.

**Table 1.** Baseline characteristics of 52 participants^†^

| Demographics |  |
| --- | --- |
| Age – years | 40.5 [28.5, 49.5] |
| Sex – no. (%) |  |
| Female | 44 (84.6) |
| Male | 8 (15.4) |
| Body mass index<sup>‡</sup>– kg/m<sup>2</sup> | 20.9 [18.7, 22.9] |
| Smoking habit – no. (current / past / never) | 0 / 6 / 46 |
| Alcohol drinking habit – no. (daily / often / never) | 7 / 30 / 15 |
| Allergies – no. (%) | 9 (17.3) |
| Comorbidities |  |
| Number of comorbidities – no. | 0 [0, 1] |
| Hypertension – no. (%) | 6 (11.5) |
| Diabetes – no. (%) | 3 (5.8) |
| Dyslipidemia – no. (%) | 5 (9.6) |
| Hyperuricemia – no. (%) | 1 (1.9) |
| Coronary heart disease – no. (%) | 0 (0.0) |
| Arrhythmia – no. (%) | 3 (5.8) |
| Stroke – no. (%) | 0 (0.0) |
| Lung disease – no. (%) | 4 (7.7) |
| Thyroid disease – no. (%) | 1 (1.9) |
| Atopic dermatitis – no. (%) | 8 (15.4) |
| Autoimmune disease – no. (%) | 1 (1.9) |
| Cancer – no. (%) | 1 (1.9) |

<sup>†</sup> Continuous variables are presented as median [1<sup>st</sup> quartile, 3<sup>rd</sup> quartile], and categorical variables are presented as number (%).
<sup>‡</sup> Body mass index was calculated using the following equation: body weight (kg) / height (m) / height (m).

### Dynamics of anti-spike IgG titers

The dynamics in anti-spike IgG titer are shown in Figure. Black circles and gray lines indicate anti-spike IgG titers and their dynamics of each participant, while orange squares and lines represent the GMT of anti-spike IgG and their dynamics of whole study participants. One participant was excluded from the result of two months after the third vaccination because the anti-nucleocapsid IgM was positive at two months after the third vaccination. After the first vaccination, all participants had an anti-spike IgG level ≥ 50 AU/ml and the GMT three weeks after the first vaccination (just before the second vaccination) was 1024.3 AU/ml. The GMT of anti-spike IgG increased to 10361 AU/ml one month after the second vaccination. The anti-spike IgG peaked at one month after the second vaccination and continued to decrease until eight months after the vaccination (just before the third vaccination). The GMT of anti-spike IgG at eight months after the second vaccination was 578.6 AU/ml, representing an average decrease rate of 93.5% from one month after the second vaccination. One month after the third vaccination, the anti-spike IgG titer of each participants increased an average of 4450% compared to just before the third vaccination and an average of 191% compared to one month after the second vaccination; the GMT of anti-spike IgG increased to 17400 AU/ml. Same as after the second vaccination, anti-spike IgG levels declined two months compared to one month after the third vaccination, with the GMT of anti-spike IgG 11185 AU/ml. However, the average decline rate was 35.7%, a significant slower pace of decline than the 59.1% rate of decline after the second vaccination (*P* < 0.01).

### Systemic adverse reactions and laboratory changes

Table 2 shows the differences of systemic adverse reactions after the second and third vaccinations. After both the second and third vaccinations, approximately half of the participants experienced fever; more participants had a fever of 38 °C or higher after the third vaccination than the second, but there was no statistical difference. The proportion experiencing fatigue also did not differ between the second and third vaccinations, with approximately 60% experiencing fatigue at either time. On the other hand, headache and axillary lymphadenopathy were more common after the third vaccination. After the third vaccination, headache was twice and axillary lymphadenopathy was seven times higher as common as after the second vaccination.

**Table 2.**
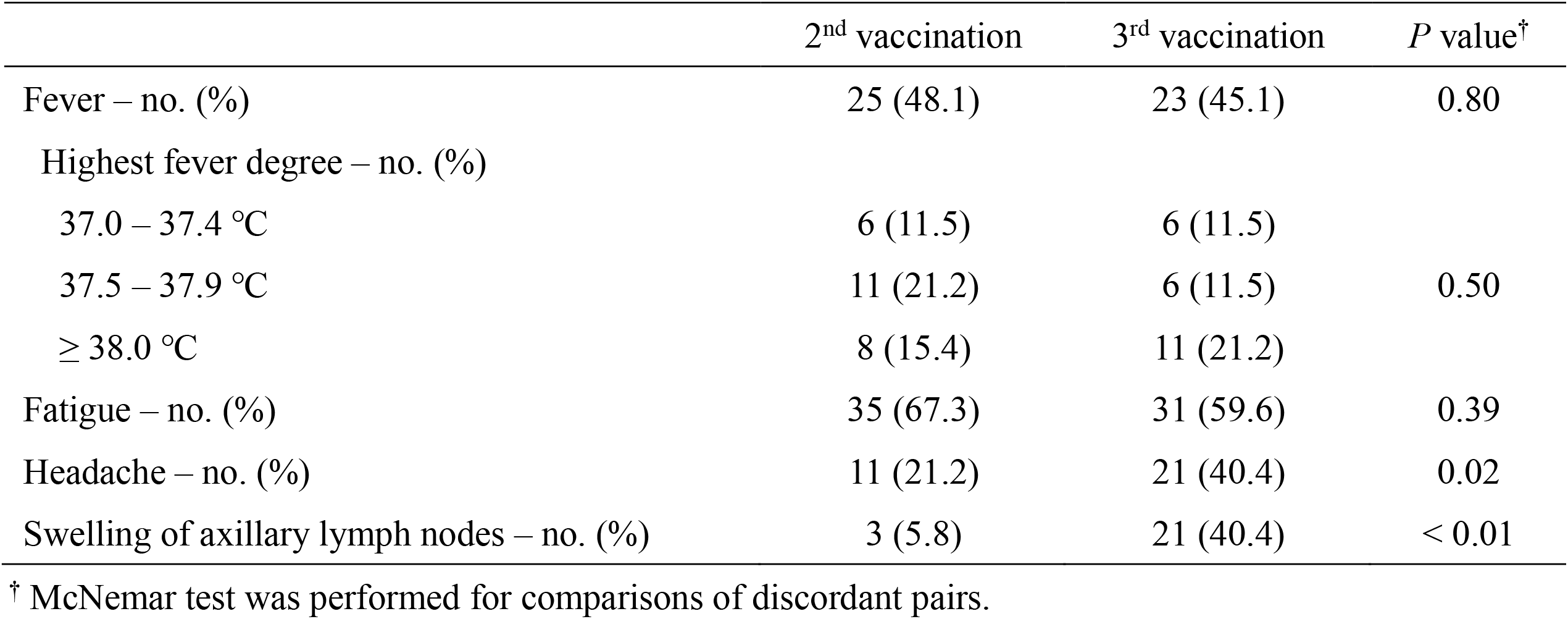
Comparison of adverse reactions at the 2^nd^ and 3^rd^ vaccinations

Table 3 shows the laboratory change between before and after each vaccination. In this study, we measured total bilirubin, AST, ALT, γ-GTP, and serum creatinine along with the anti-spike IgG. Estimated glomerular filtration rates were calculated using the Cockcroft-Gault formula. Blood tests before the first vaccination were within normal limits for most of the study participants. Blood test values hardly changed before and after each vaccination. Values of ALT and γ-GTP decreased by 1 IU/ml after the first vaccination, but we do not consider this to be of morbid significance.

**Table 3.** Comparison of the laboratory data before and after the vaccinations^†^

|  | Before vaccination | After vaccination<sup>‡</sup> | Change of value | P value<sup>§</sup> |
| --- | --- | --- | --- | --- |
| **1<sup>st</sup> vaccination** |  |  |  |  |
| Total bilirubin – mg/dl | 0.6 [0.5, 0.9] | 0.6 [0.4, 0.8] | 0.0 [-0.2, 0.0] | 0.15 |
| Aspartate aminotransferase – IU/l | 20 [16, 23] | 19 [17, 21] | 0.0 [-1.5, 1.0] | 0.57 |
| Alanine aminotransferase – IU/l | 15 [12, 20] | 14 [11, 19] | -1 [-3, 1] | 0.046 |
| γ-glutamyl transpeptidase – IU/l | 16 [14, 25] | 16 [13, 28] | -1 [-3, 0] | 0.01 |
| Serum creatinine – mg/dl | 0.63 [0.57, 0.70] | 0.62 [0.56, 0.69] | 0.00 [-0.03, 0.05] | 0.46 |
| Estimated glomerular filtration rate – ml/min/m<sup>2</sup> | 89.2 [78.6, 99.9] | 91.3 [76.0, 97.4] | 0.0 [-6.3, 5.1] | 0.57 |
| **2<sup>nd</sup> vaccination** |  |  |  |  |
| Total bilirubin – mg/dl | 0.6 [0.4, 0.8] | 0.6 [0.5, 0.8] | 0.0 [-0.1, 0.1] | 0.25 |
| Aspartate aminotransferase – IU/l | 19 [17, 21] | 18 [16, 21] | 0 [-2, 1] | 0.43 |
| Alanine aminotransferase – IU/l | 14 [11, 19] | 14 [11, 19] | 0 [-2, 2] | 0.94 |
| γ-glutamyl transpeptidase – IU/l | 16 [13, 28] | 17 [13, 28] | 0 [-1, 1] | 0.93 |
| Serum creatinine – mg/dl | 0.62 [0.56, 0.69] | 0.61 [0.55, 0.68] | 0.00 [-0.05, 0.02] | 0.41 |
| Estimated glomerular filtration rate – ml/min/m<sup>2</sup> | 91.3 [76.0, 97.4] | 90.4 [74.7, 101.1] | 0.0 [-3.5, 6.1] | 0.43 |
| **3<sup>rd</sup> vaccination** |  |  |  |  |
| Total bilirubin – mg/dl | 0.6 [0.5, 0.8] | 0.6 [0.5, 0.8] | 0.0 [-0.1, 0.1] | 0.46 |
| Aspartate aminotransferase – IU/l | 18 [16, 21] | 19 [17, 22] | 0 [-2, 2] | 0.30 |
| Alanine aminotransferase – IU/l | 14 [11, 19] | 16 [11, 21] | 0 [-1, 2] | 0.47 |
| γ-glutamyl transpeptidase – IU/l | 15 [13, 20] | 16 [14, 23] | 0 [-1, 2] | 0.10 |
| Serum creatinine – mg/dl | 0.62 [0.57, 0.69] | 0.62 [0.55, 0.70] | 0.00 [-0.04, 0.03] | 0.50 |
| Estimated glomerular filtration rate – ml/min/m<sup>2</sup> | 90.4 [80.8, 98.1] | 89.2 [74.8, 101.3] | 0.0 [-3.8, 6.9] | 0.44 |

<sup>†</sup> Data are shown as median [1<sup>st</sup> quartile, 3<sup>rd</sup> quartile].
<sup>‡</sup> Blood samples were collected three weeks after the 1<sup>st</sup> vaccination (just before the 2<sup>nd</sup> vaccination), one month after the 2<sup>nd</sup> and 3<sup>rd</sup> vaccinations.
<sup>§</sup> P values were calculated using Wilcoxon signed-rank sum test.
<sup>||</sup> Parameter was calculated using the following equation: $194 * \text{serum creatinine}^{-1.094} * \text{age}^{-0.287} (* 0.739 \text{ [if women]})$ .

### Factors influence on the anti-spike IgG titer after the third vaccination

We examined factors affecting the anti-spike IgG titer, such as age, BMI, sex, habits, comorbidities, and systemic adverse reactions to the vaccination. As we previously reported age was the most substantial factor affecting the anti-spike IgG titer induced by the vaccination, there was a significantly negative association between age and the anti-spike IgG titer one month after the third vaccination (*r* = -0.31, *P* = 0.02). However, this association disappeared two months after the third vaccination (*r* = -0.22, *P* = 0.13). BMI, sex, smoking habits, and any comorbidities did not correlate with anti-spike IgG titers both one and two months after the third vaccination. Although we previously reported that daily alcohol drinkers had significantly lower anti-spike IgG titer, there was no significant association between alcohol drink habits and anti-spike IgG titer after the third vaccination in this study. Regarding systemic adverse reactions, fever and fatigue did not correlate with anti-spike IgG titer; however, those who experienced headache or axillary lymphadenopathy tended to have higher anti-spike IgG titer after the third vaccination. Using antipyretics also did not affect the anti-spike IgG titer. After adjustment for age, only those who experienced a headache at the third vaccination had significantly higher anti-spike IgG titer one month after the third vaccination (29760 vs. 22182 AU/ml, respectively, *P* = 0.04).

We also examined the associations between anti-spike IgG titers before and after the third vaccination. The participants were divided into two groups by anti-spike IgG titer one month after the second vaccination. Participants with anti-spike IgG titers below 10000 AU/ml were grouped into the lower group, and those with anti-spike IgG titers greater than 10000 AU/ml were grouped into the higher group. As expected, the higher group always had significantly higher anti-spike IgG titers than the lower group after the second vaccination. Although the GMT of anti-spike IgG in the higher group was about three times that of the lower group one month after the second vaccination (17458 vs. 6148.8 AU/ml), the difference just before the third vaccination (eight months after the second vaccination) was about 1.7 times (732.1 vs. 438.1 AU/ml), indicating the difference between the two groups had narrowed. One month after the third vaccination, the ant-spike IgG titer of both groups increased by a factor of about 30; the GMT of the higher group and the lower group were 22067 and 11851 AU/ml, respectively. Compared with one month after the second vaccination, the GMT of the higher group increased by a factor of 1.15, but that of the lower group increased by a factor of 1.87, indicating that the third vaccination might be more effective for the lower group than the higher group.

## Discussion

The following three findings emerged from this prospective observational study. First, anti-spike IgG titers increased considerably after the third vaccination, and the GMTs of anti-spike IgG after the third vaccination were higher than those after the second vaccination. In addition, the decline rate in anti-spike IgG after the third vaccination was slower than after the second vaccination, suggesting that the vaccine-induced anti-spike IgG titer may be retained for a more extended period. Second, participants with relatively low anti-spike IgG titers after the second vaccination had a higher rate of increase in anti-spike IgG after the third vaccination than those with relatively high anti-spike IgG titers. As a result, the GMT of anti-spike IgG of study participants increased, with less differences between participants. Our results indicate that the third dose is more effective for those with low immunogenic to the primary immunization. Finally, no severe adverse reactions were observed for the series of vaccination, including the third dose, indicating that the BNT162b2 vaccine appeared to be well tolerated.

In December 2021, Israel became the first country in the world to launch a fourth dose of vaccination. A study of healthcare workers in Israel who received a fourth dose of BNT162b2 or mRNA-1273 four months after the third dose in a series of three BNT162b2 doses showed that IgG titers increased 3 to 4 times before the fourth dose [14]. However, the IgG titers after the fourth vaccination were just slightly higher than those after the third vaccination. The results of this study suggest that there may be an upper limit to the immunity induced by the mRNA vaccines and that the third vaccination may have achieved maximal immunogenicity. In our study, anti-spike IgG titers increased with the third vaccination by a factor of 4.5 compared to those before the third vaccination and by a factor of 1.9 compared to those after the second vaccination. In addition, participants with relatively low anti-spike IgG titers after the second vaccination had a higher rate of increase in anti-spike IgG titers after the third vaccination than those with relatively high anti-spike IgG titers. According to our result, the GMT of anti-spike IgG of our study participants increased, with less differences between groups. Along with previous studies [19-21], our findings also suggest that the third vaccination restored the vaccine effectiveness against SARS-CoV-2 disease, but mRNA vaccine immunogenicity may have an upper limit.

Vaccination is thought to be the most effective method of preventing infectious diseases, and there are several different types of vaccines. Attenuated vaccines use a weakened or attenuated form of the germ. Because attenuated vaccines are similar to natural infection, just one or two vaccine doses can create a strong and long-lasting immune response. Inactivated vaccines contain the inactivated micro-organisms destroyed by chemicals, heat, or radiation. Therefore, inactivated vaccines do not provide immunity as strong as attenuated vaccines, and several vaccine doses over time may need to obtain adequate immunity against diseases. Subunit, recombinant, polysaccharide, and conjugate vaccines use specific pieces of germ, such as its protein or capsid. Although these vaccines induce a robust immune response to the targeted parts of the germ, booster shots are necessary to obtain ongoing protection against diseases. Since mRNA vaccines were first used in human beings, how immunogenic they are and how many doses are required to provide adequate immunity is unclear. In our current and previous reports [16], the GMT of anti-spike IgG declined to approximately 40% from one month to two months after the second vaccination. In the current study, we found that the decline rate of the GMT became significantly slower after the third vaccination. Our results suggest that mRNA vaccines require booster vaccinations to induce adequate immunity, like inactivated vaccines such as the hepatitis B vaccine.

Fever, headache, fatigue, and pain at the injection site were the most known adverse reactions to COVID-19 vaccines, and overall, most adverse reactions were mild and short-lived [22, 23]. Although very rare, there have been reports of serious adverse reactions. The following four serious adverse reactions to certain types of COVID-19 vaccination have been found; anaphylaxis, thrombosis with thrombocytopenia syndrome, myocarditis and pericarditis, and Guillain-Barré syndrome [24-26]. There are also some reports of death after the COVID-19 vaccinations. According to the Vaccine Adverse Event Reporting System in the United States, preliminary death rates after the COVID-19 vaccination were 0.0024% [27]. Along with these previous reports, BNT162b2 appears to be well tolerated in this study. Even though at the third dose, blood tests showed hepatic enzymes or serum creatinine did not change. Regarding systemic reactions, the rates of fever and fatigue did not differ between the second and third vaccinations. Although headache and axillary lymphadenopathy were more observed with the third vaccination than with the second vaccination, no severe adverse reactions were observed.

Limitations in this study should be noted. Because the anti-spike IgG titer measured in this study is against the original strain of SARS-CoV-2, it is difficult to estimate the vaccine efficacy against the omicron variant. The observed anti-spike IgG titer might be estimated lower against the omicron variant. Moreover, we did not assess cell-mediated immunity. However, a higher anti-spike IgG titer is still considered protective against SARS-CoV-2 infection, including the omicron variant. Therefore, our results suggest that the third vaccination might restore the vaccine effectiveness against SARAS-CoV-2 infection. In addition, the number of participants was small, and participants were young. Additional research with many participants and a more comprehensive age range is necessary to validate our findings. Finally, we could not assess the effect of the third dose of vaccine on COVID-19 prevention because there was only one participant possibly infected with SRAS-CoV-2, and there was no control group. Further analyses with nationwide surveys are necessary to confirm the efficacy of the additional COVID-19 vaccination.

In conclusions, the third dose of BNT162b2 vaccination successfully increased anti-spike IgG titer and the efficacy of vaccination might be maintained longer because the decline rate of anti-spike IgG is slower than after the second vaccination. In addition, the third vaccination may be more effective for those with low immunogenic to the primary immunization.

## Data Availability

All data produced in the present study are available upon reasonable request to the authors.

## Abbreviations

COVID-19: coronavirus disease 2019
mRNA: messenger RNA
SARS-CoV-2: severe acute respiratory syndrome coronavirus 2
IgG: immunoglobulin G
AU/ml: arbitrary units per milliliter
IgM: immunoglobulin M
AST: aspartate aminotransferase
ALT: alanine aminotransferase
γ-GTP: γ-glutamyl transpeptidase
GMT: geometric mean titer

## Acknowledgments

The authors thank Ms. Ryoko Nakashima for managing the dataset and Drs. Kahori Miyoshi, Yuichi Hara, Jun Hayashi, and Hiroshi Hara for scientific advice.

## Figure Legends

**Figure.**
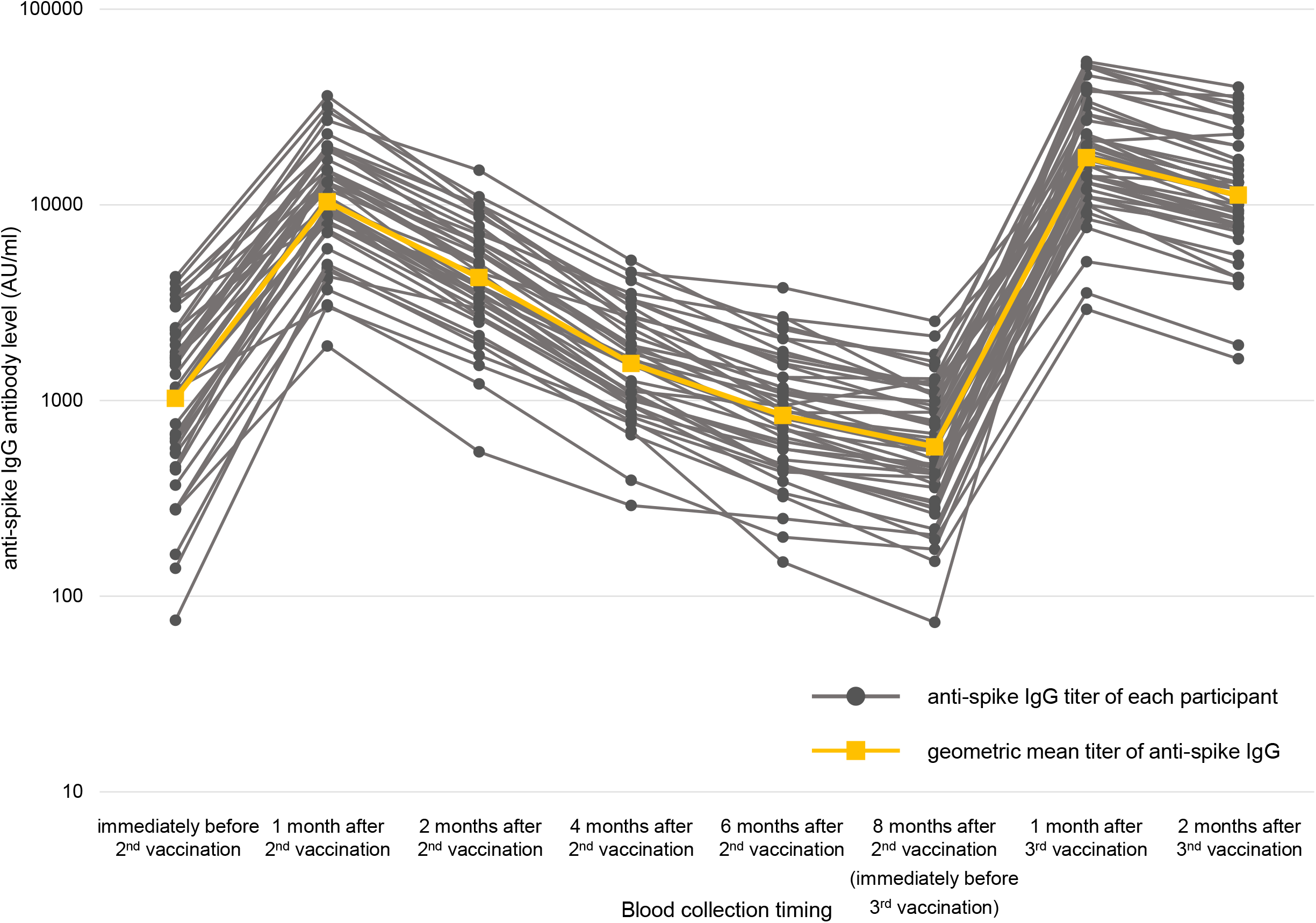
Dynamics of anti-spike IgG titers. Dynamics of anti-spike IgG titer of each participants (black circles and gray lines) and geometric mean titer of anti-spike IgG (orange squares and lines) are shown. Compared to one month after the second vaccination, the geometric mean titer of anti-spike IgG before the third vaccination decreased by about one-tenth. One month after the third vaccination, the geometric mean titer of anti-spike IgG increased 30-fold before the third vaccination and 1.7-fold one month after the second vaccination.

